# A double-blind sham-controlled phase 1 clinical trial of tDCS of the dorsolateral prefrontal cortex in cocaine inpatients: craving, sleepiness, and contemplation to change

**DOI:** 10.1101/2020.10.09.20209676

**Authors:** Pierre-Olivier Gaudreault, Akarsh Sharma, Abhishek Datta, Ester M. Nakamura-Palacios, Sarah King, Pias Malaker, Ariella Wagner, Devarshi Vasa, Muhammad A. Parvaz, Lucas C. Parra, Nelly Alia-Klein, Rita Z. Goldstein

## Abstract

Impaired inhibitory control accompanied by enhanced salience attributed to drug-related cues, both dorsolateral prefrontal cortex (dlPFC) functions, are hallmarks of drug addiction, contributing to worse symptomatology including craving. dlPFC modulation with transcranial direct current stimulation (tDCS) showed craving reduction in inpatients with cocaine use disorder (CUD). Our study aimed at assessing feasibility of a longer tDCS protocol in CUD (15 vs. the common five/10 sessions), and replicability of previous results.

In a randomized double-blind sham-controlled protocol, 17 inpatients were assigned to either a real-tDCS (right anodal/left cathodal) or a sham-tDCS condition, for 15 sessions. Primary outcome measures were self-reported craving, anxiety, depression, and quality of life. Secondary measures included sleepiness, readiness to change drug use, and affect. We also assessed cognitive function including impulsivity.

An 82% retention rate demonstrated feasibility. Partially supporting previous results, there was a trend for self-reported craving to decrease in the real-tDCS group more than the sham group, an effect that would reach significance with 15 subjects per group. Quality of life and impulsivity improved over time of treatment in both groups. Significant group × time interactions showed improvements after treatment only in the real-tDCS group for daytime sleepiness and readiness to change drug use. One-month follow-up suggested transient effects of tDCS on sleepiness and craving.

This study suggests that more subjects are needed to show a unique effect of real-tDCS on craving and to examine the duration of effect. Increased vigilance and motivation to change in the real-tDCS group suggest fortification of dlPFC-supported executive functions.

## Introduction

Drug addiction is a chronic brain disorder characterized by a relapsing cycle of compulsive drug seeking and using followed by withdrawal and craving that repeat despite harmful consequences (Volkow & Fowler, 2000). The underlying mechanisms encompass impairments in prefrontal cortex-mediated functions that include reduced inhibitory control of automatic or maladaptive behaviors, as well as the attribution of excessive salience to the drug and drug-related cues at the expense of non-drug related stimuli and cues, in a syndrome referred to as the impaired response inhibition and salience attribution (iRISA; (Goldstein & Volkow, 2002, 2011). Specifically, dysfunction in dorsolateral subregions of the prefrontal cortex (dlPFC), associated with impairments in executive control, is thought to contribute to impulsive, compulsive, inflexible, or stimulus-driven behaviors (Lubman *et al.*, 2004; Simon *et al.*, 2007; Garavan *et al.*, 2008), including craving. Drug craving, the intense subjective urge to use drugs, predicts clinical outcomes such as drug use and the propensity to relapse in drug addicted individuals (Ludwig & Wikler, 1974; Weiss *et al.*, 2003; Paliwal *et al.*, 2008; Chaves *et al.*, 2011). Craving has therefore been considered an important therapeutic target in the treatment of substance use disorders (Tiffany & Wray, 2012; Tiffany *et al.*, 2012). However, as most pharmacological treatments and behavioral therapies for this purpose show inconsistent results (Hennessy *et al.*, 2003; Sofuoglu & Kosten, 2006; Zilverstand *et al.*, 2016; Mellentin *et al.*, 2017; Liu & Li, 2018), reducing craving in drug addiction remains a challenge.

Through the use of magnetic or electric brain stimulation, neuromodulation has recently gained momentum as an adjunct therapy for various psychiatric conditions including bipolar disorder and depression as well as for pain management (Kuo *et al.*, 2014; Rapinesi *et al.*, 2015; Sampaio-Junior *et al.*, 2018). By targeting specific brain regions implicated in addiction such as the dlPFC, neuromodulation has also been used in drug addicted individuals to reduce craving and maladaptive addiction-related behaviors (see Ekhtiari *et al.*, 2019 for review). A recent meta-analysis of studies using non-invasive brain stimulation over the dlPFC, the medial prefrontal cortex, and the anterior cingulate cortex, showed a specific strong main effect on drug craving in stimulant-addicted individuals (Ma *et al.*, 2019). For instance, repeated sessions of transcranial magnetic stimulation (TMS), which consists of focal electromagnetic pulses that elicit neuronal firing in the targeted region under a coil positioned on the scalp (Hallett, 2007; Wagner *et al.*, 2007; Rossi *et al.*, 2009), showed reductions in subjective craving as well as drug intake when the dlPFC was targeted in cocaine addicted individuals (Bolloni *et al.*, 2016; Rapinesi *et al.*, 2016; Terraneo *et al.*, 2016). A study in young males with cocaine use disorder (CUD) found that even a single session of repetitive TMS had an effect (albeit transient) on reducing craving (Camprodon *et al.*, 2007). Although these studies show promising outcomes, the widespread use of TMS is limited by several factors (e.g., its lack of portability, the expertise required for administration, expense, etc.). As a battery-powered portable device that can be used remotely and repetitively, transcranial direct current stimulation (tDCS) directly applies small amounts of continuous low-intensity electric currents to the scalp in order to facilitate, in a polarity-dependent fashion, cortical excitability through changes in neuronal membrane potentials (Bindman *et al.*, 1964; Nitsche & Paulus, 2000). The repeated use of tDCS specifically targeting the dlPFC either daily or every other day for up to 10 days showed a reduction in subjective self-reported craving (Batista *et al.*, 2015; de Almeida Ramos *et al.*, 2016; Klauss *et al.*, 2018), and a decrease in electrophysiological measures of drug-related cue reactivity (Conti & Nakamura-Palacios, 2014; Conti *et al.*, 2014; Nakamura-Palacios *et al.*, 2016) as well as in risk-taking behaviors (Gorini *et al.*, 2014) in individuals with CUD (studies mostly conducted in Brazil). Recently, a study combining tDCS and positron emission tomography in healthy individuals suggested that repeated tDCS over the dlPFC induced dopamine release in the ventral striatum, a hub within the reward and motivation network, pointing to a possible putative neurochemical mechanism of such scalp-based brain stimulation (Fonteneau *et al.*, 2018).

The goal of the current Phase 1 clinical trial was therefore to replicate the previous tDCS findings in treatment-seeking individuals with CUD living in an inpatient drug-rehabilitation facility in a northeastern US urban area. Given an ultimate goal of developing a home-based self-administration protocol, a related main aim was to assess the feasibility of administering 15 repeated sessions of tDCS in contrast to the previous shorter tDCS administration protocols. Based on the previous findings (Batista *et al.*, 2015), our main hypothesis was that real-tDCS will be associated with a decrease in the subjective reports of cocaine craving as well as symptoms of anxiety and depression, and with an increase in quality of life measures. In line with the literature, we also assessed the effect of repeated tDCS on secondary outcome measures, including subjective sleep measures (Minichino *et al.*, 2014; Frase *et al.*, 2016; Sheng *et al.*, 2018; Charest *et al.*, 2019), readiness to change (Opsal *et al.*, 2019), and positive and negative affect (Wiegand *et al.*, 2019). Neuropsychological tests measuring verbal fluency (including drug-related fluency as a proxy of cue-reactivity), temporal discounting/impulsivity, and self-reported sensitivity to reinforcement of rewards (Goldstein *et al.*, 2010), were also included for exploratory purposes.

## Materials and Methods

### Participants

Twenty-nine detoxified and abstinent individuals with CUD between the ages of 18 and 65 years were recruited from, and studied at, an inpatient addiction treatment facility (Samaritan Daytop Village – Jamaica, Queens, NY). Potential participants were identified from rolling admissions and were screened for the study following a period of detoxification (≥two weeks since last drug use). All participants met the diagnostic criteria for CUD as corroborated by a comprehensive diagnostic interview that included the Structured Clinical Interview for *Diagnostic and Statistical Manual of Mental Disorders*, Fifth Edition (First, 2015) and the Addiction Severity Index (McLellan *et al.*, 1992) to evaluate lifetime drug use and recency of drug use. Drug use questionnaires and scales also included the 5-item Severity of Dependence Scale (Foltin, 1999), the 18-item Cocaine Selective Severity Assessment (Kampman *et al.*, 1998) for withdrawal symptoms within 24 hours of the interview, and the Cocaine Negative Consequences Checklist (Michalec *et al.*, 1996). Exclusion criteria were: 1) current or past history of a major neurological disorder, seizures or a neurodevelopmental disorder; 2) a history of head trauma with loss of conciousness (for ≥30 minutes); 3) history of Axis I disorders (with the exception of CUD) associated with psychotic symptoms; 4) medication use with known effects on the central nervous system within the past 6 months (except psychotropics for symptoms of depression/anxiety/post traumatic stress disorder); 5) a clinically significant and/or unstable medical illness or infection; 6) the presence of contraindicated metallic implants or devices that may be affected by electrical stimulation; and 7) pregnancy or breast feeding in women. All participants provided full written consent in accordance with both the Icahn School of Medicine at Mount Sinai and the Samaritan Daytop Village’s institutional review boards. This study was registered under Clinical Trials.gov number NCT03833583.

Of the 29 inpatients with CUD who were consented, one was withdrawn from the study (due to inability to attend the appointments) (Figure 1). Of the remaining 28 subjects, 17 individuals qualified for the study (meeting the inclusion/exclusion criteria), completing the pre-treatment behavioral assessments before being randomly assigned to either the real-tDCS or the sham-tDCS condition (control group). During the course of treatment, three participants withdrew from the study, all in the sham-tDCS group (this intriguing difference between the groups in drop-out rate should be further explored with larger samples). They were considered lost to follow-up: two relapsed (after 4 and 9 sessions, respectively) leaving the treatment facility on their own accord; the third participant was transferred to another facility (that did not allow participation in research) after 12 sessions. Of the 14 subjects who completed all 15 tDCS sessions, 12 participants (real-tDCS: N = 7; sham-tDCS: N = 5) attended a one-month follow-up visit (2 participants were not able to attend due to a change in treatment location). All subjects continued taking their prescribed medications throughout the study. The demographic, neuropsychological and drug-related variables of the subjects who completed all 15 tDCS sessions in both groups are presented in Table 1.

**Figure 1:**
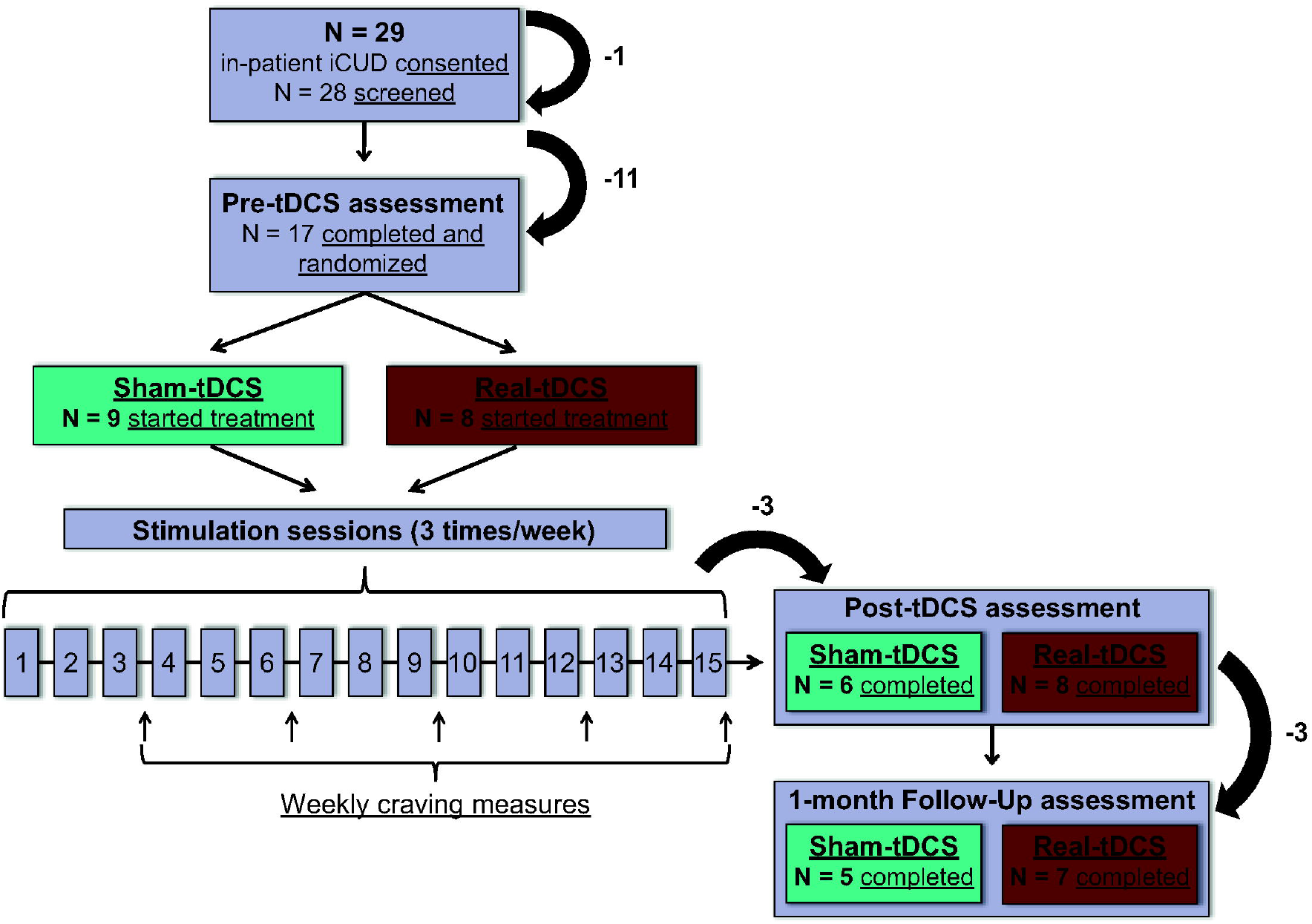
Representation of Phase 1 clinical trial protocol and recruitment, randomization and assessment of participants.

**Table 1.** Demographic, neuropsychological, clinical drug use, and outcome measures in sham- and real-tDCS groups.

|  | Sham-tDCS (N = 6) |  | Real-tDCS (N = 8) |  | Main effects (Cohen's *d*) |  |  |
| --- | --- | --- | --- | --- | --- | --- | --- |
|  | Pre-sham | Post-sham | Pre-tDCS | Post-tDCS | Group | Time | Interaction |
| *Demographics* |  |  |  |  |  |  |  |
| Age (years) | 46.7 ± 13.9 | -- | 40.4 ± 10.2 | -- | $t_{12} = 1.0$ | N/A | N/A |
| Sex (male/female) | 6 / 0 | -- | 6 / 2 | -- | $\chi^2 = 1.8$ | N/A | N/A |
| Race (African-American/Caucasian/ Mixed) | 3 / 3 / 0 | -- | 1 / 6 / 1 | -- | $\chi^2 = 2.8$ | N/A | N/A |
| Ethnicity (Hispanic/Non-Hispanic) | 2 / 4 | -- | 3 / 5 | -- | $\chi^2 = 0.0$ | N/A | N/A |
| Education (years) | 11.1 ± 1.4 | -- | 12.3 ± 2.6 | -- | $t_{12} = -1.0$ | N/A | N/A |
| Handedness (Right/Left/Ambidextrous) | 6 / 0 / 0 | -- | 6 / 0 / 1<sup>a</sup> | -- | $\chi^2 = 0.9$ | N/A | N/A |
| *Neuropsychological tests* |  |  |  |  |  |  |  |
| WRAT – Reading Scale (standard score) | 98.5 ± 13.5 | -- | 92.0 ± 10.3<sup>a</sup> | -- | $t_{11} = 1.0$ | N/A | N/A |
| WASI – Matrix Reasoning (scaled score) | 9.8 ± 3.3 | -- | 9.0 ± 2.0<sup>a</sup> | -- | $t_{11} = 0.6$ | N/A | N/A |
| Semantic (neutral) verbal fluency | 17.6 ± 2.0 | 20.3 ± 1.5 | 18.0 ± 1.7 | 17.8 ± 1.3 | $F = 0.3 (d = .30)$ | $F = 1.3 (d = .64)$ | $F = 1.6 (d = .74)$ |
| Drug verbal fluency | 16.8 ± 1.9 | 17.0 ± 1.6 | 16.8 ± 1.6 | 17.1 ± 1.4 | $F = 0.0 (d = .00)$ | $F = 0.2 (d = .24)$ | $F = 0.0 (d = .09)$ |
| Kirby delay discounting task | 0.13 ± 0.04 | 0.07 ± 0.02 | 0.12 ± 0.03 | 0.06 ± 0.01 | $F = 0.2 (d = .24)$ | **$F = 5.7 (d = 1.4)^*$** | $F = 0.0 (d = .10)$ |
| *Clinical drug use measures* |  |  |  |  |  |  |  |
| Cocaine use lifetime (years) | 14.3 ± 9.0 | -- | 8.1 ± 7.6 | -- | $t_{12} = 1.4$ | N/A | N/A |
| Duration of current abstinence (days) | 46.2 ± 13.4 | 77.0 ± 15.4 | 59.4 ± 11.6 | 84.6 ± 13.3 | $F = 0.3 (d = .33)$ | **$F = 31.5 (d = 3.2)^{**}$** | $F = 0.3 (d = .32)$ |
| Severity of dependence scale | 10.8 ± 3.3 | -- | 11.0 ± 2.4 | -- | $t_{12} = -0.1$ | N/A | N/A |
| Cocaine selective severity assessment | 17.8 ± 5.0 | 20.3 ± 5.3 | 14.9 ± 4.4 | 9.0 ± 4.6 | $F = 1.4 (d = .68)$ | $F = 0.3 (d = .30)$ | $F = 1.7 (d = .75)$ |
| Cocaine negative consequences checklist | 43.3 ± 6.2 | 49.2 ± 6.1 | 30.9 ± 5.4 | 29.3 ± 5.3 | **$F = 6.5 (d = 1.5)^*$** | $F = 0.2 (d = .24)$ | $F = 0.5 (d = .42)$ |
| *Primary outcome measures* |  |  |  |  |  |  |  |
| 5-item OCCS craving subscale | 5.5 ± 1.6 | 5.8 ± 1.5 | 4.4 ± 1.3 | 1.6 ± 1.3 | $F = 3.1 (d = 1.0)$ | $F = 0.9 (d = .53)$ | $F = 1.4 (d = .68)$ |
| 5-item cocaine cravings | 14.2 ± 4.6 | 7.3 ± 3.5 | 11.1 ± 4.0 | 6.9 ± 3.0 | $F = 0.1 (d = .22)$ | **$F = 4.2 (d = 1.2)^{\dagger}$** | $F = 0.2 (d = .28)$ |
| Hamilton anxiety rating scale | 8.0 ± 3.4 | 9.3 ± 3.8 | 6.5 ± 2.9 | 5.3 ± 3.3 | $F = 0.5 (d = .40)$ | $F = 0.0 (d = .00)$ | $F = 0.3 (d = .30)$ |
| Hamilton depression rating scale | 8.5 ± 4.0 | 4.8 ± 1.8 | 5.6 ± 3.4 | 3.3 ± 1.6 | $F = 0.4 (d = .38)$ | $F = 1.9 (d = .79)$ | $F = 0.1 (d = .17)$ |
| Quality of life – Physical health | 15.2 ± 1.2 | 16.7 ± 1.2 | 15.5 ± 1.1 | 16.1 ± 1.0 | $F = 0.0 (d = .04)$ | $F = 2.0 (d = .82)$ | $F = 0.4 (d = .34)$ |
| Quality of life – Psychological | 15.3 ± 1.4 | 15.8 ± 1.2 | 15.5 ± 1.2 | 16.8 ± 1.0 | $F = 0.1 (d = .22)$ | $F = 1.1 (d = .61)$ | $F = 0.2 (d = .26)$ |
| Quality of life – Social relationship | 11.8 ± 1.8 | 13.2 ± 0.9 | 12.6 ± 1.5 | 12.1 ± 0.8 | $F = 0.0 (d = .05)$ | $F = 0.1 (d = .21)$ | $F = 0.6 (d = .45)$ |
| Quality of life – Environment | 12.7 ± 1.0 | 15.2 ± 1.2 | 12.4 ± 0.9 | 13.6 ± 1.0 | $F = 0.5 (d = .39)$ | **$F = 10.9 (d = 1.9)^{**}$** | $F = 1.2 (d = .64)$ |
| *Secondary outcome measures* |  |  |  |  |  |  |  |
| Epworth sleepiness scale | 7.8 ± 1.8<sup>b</sup> | 11.0 ± 1.7 | 7.1 ± 1.4 | 5.1 ± 1.3 | $F = 3.1 (d = 1.1)$ | $F = 0.3 (d = .32)$ | **$F = 5.2 (d = 1.4)^*$** |
| Pittsburgh sleep quality index | 7.4 ± 1.7<sup>b</sup> | 8.2 ± 2.0 | 3.9 ± 1.3 | 6.0 ± 1.6 | $F = 1.6 (d = .76)$ | **$F = 5.0 (d = 1.4)^*$** | $F = 1.0 (d = .61)$ |
| Contemplation ladder | 8.7 ± 0.5 | 8.3 ± 0.6 | 8.0 ± 0.4 | 9.1 ± 0.5 | $F = 0.0 (d = .06)$ | $F = 1.6 (d = .73)$ | **$F = 5.4 (d = 1.3)^*$** |
| PANAS – Positive affect | 38.7 ± 2.9 | 41.3 ± 2.5 | 41.5 ± 2.5 | 40.3 ± 2.2 | $F = 0.1 (d = .16)$ | $F = 0.2 (d = .26)$ | $F = 1.6 (d = .73)$ |
| PANAS – Negative affect | 19.8 ± 3.6 | 21.7 ± 3.6 | 17.8 ± 3.1 | 16.9 ± 3.1 | $F = 0.6 (d = .44)$ | $F = 0.1 (d = .21)$ | $F = 1.0 (d = .59)$ |

Notes: Data expressed as frequencies or means ± standard deviation (SD). <sup>a</sup>One datapoint was missing for two participants, one for WRAT – Reading scale and one for the WASI – Matrix score and handedness. <sup>b</sup>One participant was also missing the pre-tDCS sleepiness questionnaires. OCCS: Obsessive-Compulsive Cocaine Scale PANAS: Positive and Negative Affect Schedule; WASI: Wechsler Abbreviated Scale of Intelligence; WRAT: Wide Range Achievement Test. <sup>†</sup>trend at $p < 0.1$ ; \* $p < 0.05$ ; \*\* $p < 0.01$ .

### Double-Blind Study Procedures

Fourteen participants underwent all 15 sessions of either real-tDCS or sham-tDCS every other day over five weeks. The stimulations were administered using a battery-driven 1×1 (one anode and one cathode) portable tDCS mini-CT device (Soterix Medical, Inc., New York, USA). Two single-use 35 cm^2^ sponge electrodes pre-soaked in a saline solution were used with the cathodal electrode placed over the left hemisphere (F3 electrode) and the anodal electrode over the right hemisphere (F4 electrode) targeting the dlPFC. The anodal (positive) and cathodal (negative) nature of the stimulation refers to the polarity of the electrode placed over the targeted area (Woods *et al.*, 2016), whereby cortical excitability is increased beneath the anode and decreased beneath the cathode (Nitsche & Paulus, 2000). While TMS induces neuronal firing directly beneath the stimulation coil (Opitz *et al.*, 2011), tDCS is thought to enhance a more diffused neuronal plasticity underneath the large electrodes on the scalp, potentially reaching broader regions of interest (Datta *et al.*, 2009; Opitz *et al.*, 2013; DaSilva *et al.*, 2015; Santos *et al.*, 2016; Huang *et al.*, 2017). Electrode placement was ensured by a premeasured strap (Soterix Medical, Inc., New York, USA), fitted to the participant’s headsize, on which the electrodes were snapped. Every electrode placement was photographed prior to each stimulation in order to assess the reliability of the positioning in-between sessions. The tDCS devices were pre-programmed to deliver real- or sham-tDCS by a co-author who was not involved in data acquisition or analyses, and both the experimenters and the participants were not aware of the applied treatment. In the real-tDCS condition, each stimulation delivered 2mA for a duration of 20 minutes. Stimulations started with a 30-second ramp up, during which the device slowly increased the amount of energy delivered to reach 2mA. A ramp down to zero occurred during the last 30 seconds of the stimulation. In order to maintain effective subject blinding, the sham-tDCS condition also included the 30 seconds ramp up from zero to 2mA followed by an immediate ramp down back to zero where the device did not deliver any current. At the end of the 20-minute sham-tDCS session, the same ramp up and ramp down was reproduced once again to mimic the tingling sensation under the electrodes usually reported by subjects receiving active stimulation (Brunoni *et al.*, 2011). The actual length of the stimulation for all sessions was recorded on the tDCS device and all recordings were verified after the study in order to ensure that each participant received the intended set amount of stimulation. The blind was only removed after the completion of the entire study inclusive of data analyses.

### Behavioral Assessment and Outcome Measures

Neuropsychological tests evaluated: 1) handedness [modified Edinburgh Handedness Inventory (Milenkovic & Dragovic, 2013)]; 2) color blindness [abbreviated version of the Ishihara color blindness test (Ishihara, 1917) with all subjects except for one of normal color vision]; 3) estimated premorbid verbal IQ [Wide Range Achievement Test (Wilkinson, 1993) – Reading scale]; 4) estimated premorbid non-verbal IQ [Wechsler Abbreviated Scale of Intelligence (Wechsler, 1999) – Matrix Reasoning Subtest]; 5) verbal fluency (semantic and drug) (Goldstein *et al.*, 2007, 2009; Lezak *et al.*, 2012); and 6) the Kirby Delay Discounting Task for temporal discounting/impulsivity (Kirby *et al.*, 1999).

The primary outcome measures, aimed at replicating previous findings (Batista *et al.*, 2015), included the: 1) a subscale of the Obsessive-Compulsive Cocaine Scale (Vorspan *et al.*, 2012) that includes 5 items (1, 2, 4, 5, and 13) known to be specificaly associated with cocaine cravings (De Wildt *et al.*, 2005); 2) 5-item Cocaine Craving Questionnaire (Tiffany *et al.*, 1993) to further measure subjective cocaine craving; 3) Hamilton Anxiety Rating Scale (Hamilton, 1959) for state anxiety; 4) Hamilton Depression Rating Scale (Hamilton, 1960) for state depression; and 5) an abbreviated version of the World Health Organization Quality of Life (Skevington *et al.*, 2004) for measures of quality of life on four different domains (physical health, psychological, social relationship, and environment).

Secondary outcome measures included the Epworth Sleepiness Scale (Johns, 1991) assessing the likelihood of falling asleep in various situations (ranging from “would never doze” to “high chance of dozing”), the Pittsburgh Sleep Quality Index (Buysse *et al.*, 1989) to assess sleep quality as well as daily interviews to assess sleep quantity (number of hours of sleep during the previous night). Included were also the Contemplation Ladder (Biener & Abrams, 1991) that evaluates participants’ willingness to change their drug use (with a score of 0 being associated with: “*No thoughts about quitting. I cannot live without drugs*,” a score of 4 being associated with: “*I sometimes think about changing the way that I use drugs, but I have not planned to change it yet*,” a score of 8 being associated with: “*I still use drugs, but I will begin to change, like cutting back on the amount of drugs that I use*,” and a score of 10 being associated with: “*I have changed my drug use and will never go back to the way I used drugs before.”)*, the Positive and Negative Affect Schedule (Watson *et al.*, 1988) assessing the positive and negative dimensions of mood, and the Sensitivity to Reinforcement of Addictive and other Primary Rewards questionnaire (STRAP-R) (Goldstein *et al.*, 2010) to assess the liking and wanting of drug and non-drug rewards.

### Statistical Analyses

A first set of statistical analyses was conducted to investigate group differences at baseline on demographic variables and the IQ measures. Continuous variables were compared using Student’s T-tests whereas chi-squared (χ^2^) tests were used for categorical variables. A second set of analyses aimed at assessing pre- vs. post-tDCS effects using 2 (Group: sham vs. real-tDCS) × 2 (Time: pre- vs. post-tDCS) analysis of variance (ANOVA) with repeated measures for variables of interest that included select neuropsychological tests, clinical drug variables as well as the primary and secondary outcome variables. Daily measures of cocaine craving and sleep quantity were assessed with two 2 (Group: sham vs. real-tDCS) × 15 (Time: all study sessions) ANOVAs with repeated measures. The STRAP-R data were analysed following a previously published procedure (Goldstein *et al.*, 2010) with a 2 (Group: sham vs. real-tDCS) × 3 (Reward: food, sex, drug) × 3 (Situation: ‘current’, ‘in general’, ‘under the influence’), × 2 (Question: ‘liking’ and ‘wanting’), × 2 (Times: pre-tDCS, post-tDCS) mixed repeated-measures ANOVA. Finally, in order to investigate any potential longer-lasting effects of tDCS stimulation, additional 2 (Group: sham vs. real-tDCS) × 3 (Time: pre-tDCS, post-tDCS, and 1-month follow-up) ANOVAs with repeated measures were carried out for the primary and secondary variables that showed any significant Time or Group × Time interactions in the first set of ANOVAs. In all ANOVAs, simple effects were analyzed to follow-up significant interactions. Greenhouse-Geisser correction was used for repeated measures ANOVAs if needed. Covariates were the demographic, neuropsychological or clinical drug use measures that differed between the groups and correlated with any of the outcome measures that showed any significant results (Pocock *et al.*, 2002). For this pilot small N study, corrections for multiple comparisons were not undertaken and results were considered significant at p < 0.05 for all outcome measures. Results with p < 0.1 are reported as showing a trend.

## Results

### Demographic Variables and Neuropsychological Tests

Supporting the randomization procedure and the creation of matched groups, there were no significant group differences at baseline in any of the demographic variables or neuropsychological tests (Table 1, which includes Cohen’s d effect sizes for the repeated measures ANOVA). A main effect for Time was significant for the Kirby Delay Discounting Task, where both groups showed decreased values of *k* from pre- to post-tDCS suggesting improvement in the ability to value delayed rewards with inpatient treatment [F(1,12) = 5.68, p = 0.035, *d =* 1.4]. There were no other Time or interaction effects that reached significance for the neuropsychological tests [F(1,12) < 1.64, p > 0.225, d < .74].

### Clinical Drug Use Measures

Our 2 × 2 ANOVAs with repeated measures showed the expected significant increase in abstinence length (days) in both groups [F(1,12) = 31.5, p = 0.01, *d* = 3.2] from pre- to post-tDCS, suggesting adherence to the inpatient treatment. A significant group effect was detected for the Cocaine Negative Consequences Checklist where the real-tDCS group reported overall less negative consequences of cocaine use than the sham-tDCS group [F(1,12) = 6.53, p = 0.025, *d =* 1.5]; since this measure did not correlate with any of the outcome measures that showed significant results (below), we did not include it as a covariate in the subsequent analyses. There were no other significant effects for any of the other drug use variables including lifetime cocaine use, severity of dependence, and withdrawal symptoms at time of study.

### Primary Outcome Measures

The 2 (Group) × 2 (Time) ANOVA with repeated measures with the 5-item Obsessive-Compulsive Cocaine Scale craving subscale yielded no significant effects. However, since a trend for a Group effect (p = 0.1, one-tailed statistical test) was observed, we explored the same ANOVA with repeated measures while correcting for baseline values. This analysis showed a trend for a significant Group × Time interaction [F(1,11) = 3.885, p = 0.074, Figure 2A] where only the real-tDCS group showed decreased self-reported craving scores after 15 sessions of stimulation. Power analysis (G*Power v3.1.9.4) suggested that 15 subjects per group would allow sufficient power (80%) for detecting this effect at the .05 significance level. The ANOVA for the 5-item Cocaine Craving Questionnaire showed a trend for a Time effect [F(1,12) = 4.17, p = 0.064, *d =* 1.2, Figure 2B] that was further supported by a closer look at the self-reported craving intensity item on the Cocaine Selective Severity Assessment, showing a significant Time effect [F(1,12) = 7.15, p = 0.020, *d =* 1.5, Figure 2C], together suggesting decreased self-reported craving from pre- to post-tDCS. Similarly, the 2 × 15 ANOVA for the daily self-report of cocaine craving revealed a trend for a Time main effect [F(14,168) = 1.63, p = 0.075, d = .20, Figure 4A], with a linear contrast reaching significance (p < 0.05) to suggest a linear decrease in craving over the 15 sessions across all subjects. A Time main effect was also significant for the World Health Organization Quality of Life, the environment scale [F(1,12) = 10.92, p = 0.006, *d =* 1.9], which increased from pre- to post-tDCS in both groups, again consistent with adherence to the inpatient treatment and the overall longer abstinence. No significant effects were observed for state depression or anxiety [F(1,12) < 1.87, p > 0.197, d < .79].

**Figure 2:**
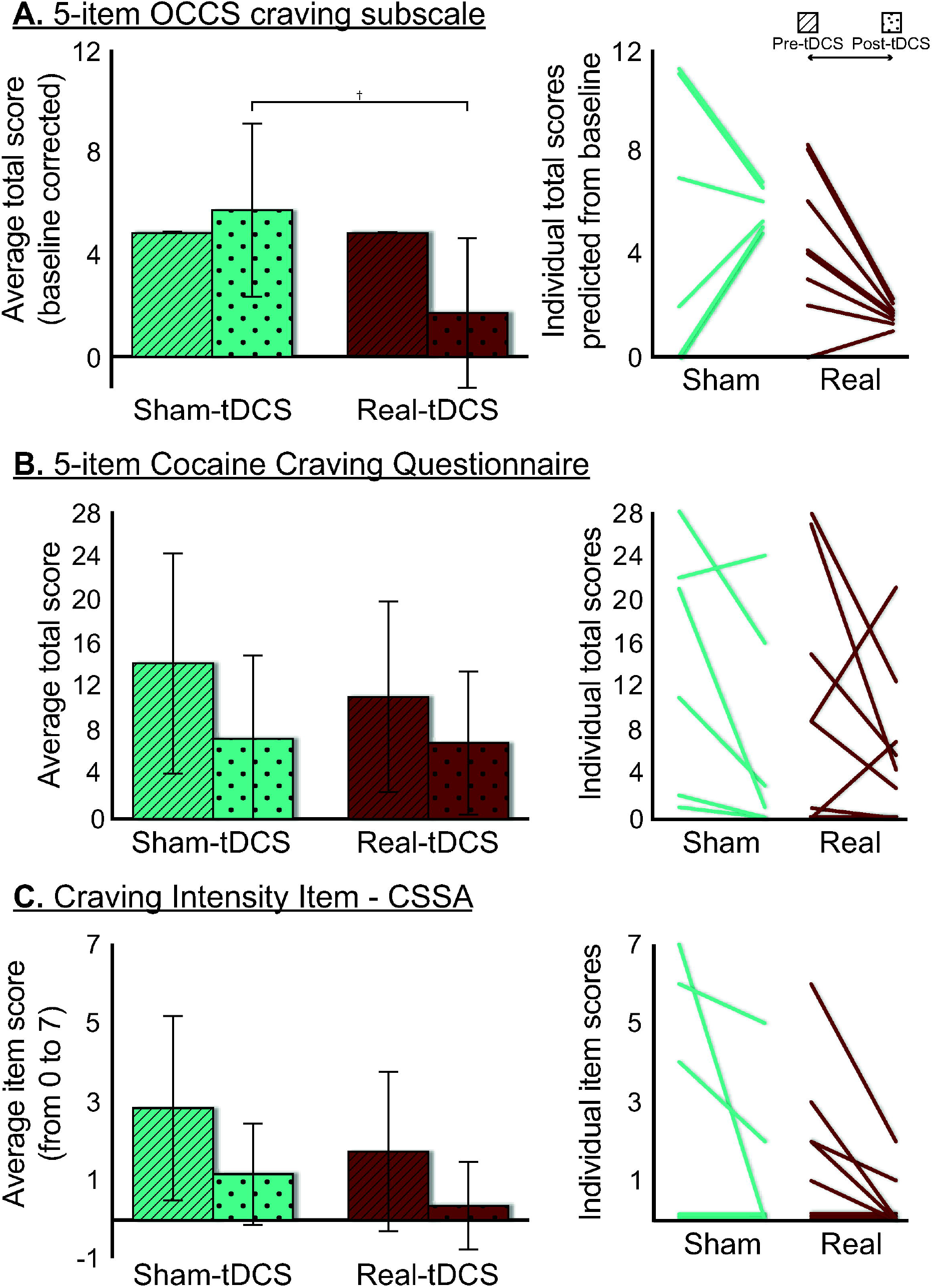
Group-averaged total scores for cocaine craving at both timepoints (pre- and post-tDCS). A) Left panel: group-averaged baseline-corrected total score on the 5-item Obsessive-Compulsive Cocaine Scale craving subscale at both study timepoints (pre- and post-tDCS). Given the trend for a Group effect (p = 0.1, one-tailed statistical test), we explored the 2 × 2 ANOVA with repeated measures while correcting for baseline values. This analysis showed a trend for a significant Group × Time interaction (^**†**^ p = 0.074) where only the real-tDCS group showed decreased self-reported craving scores after 15 sessions of stimulation. Power analysis suggested that 15 subjects per group would allow sufficient power for detecting this effect at the .05 significance level. Middle panel: Individual scores showing change from pre- to post-tDCS in each group. Right panel: All subjects scores at both study timepoints. B) Left panel: group-averaged total score on the 5-item Cocaine Craving Questionnaire at both study timepoints (pre- and post-tDCS). Results showed a trend for a Time effect (^**†**^ p = 0.064). Middle panel: Individual scores showing change from pre- to post-tDCS in each group. Right panel: All subjects scores at both study timepoints showing the trend for a Time effect. C) Left panel: group-averaged total score on the craving intensity item of the Cocaine Selective Severity Assessment at both study timepoints (pre- and post-tDCS). Results showed a significant Time effect (* p < 0.05). Middle panel: Individual scores showing change from pre- to post-tDCS in each group. Right panel: All subjects scores at both study timepoints showing the significant Time effect.

**Figure 3:**
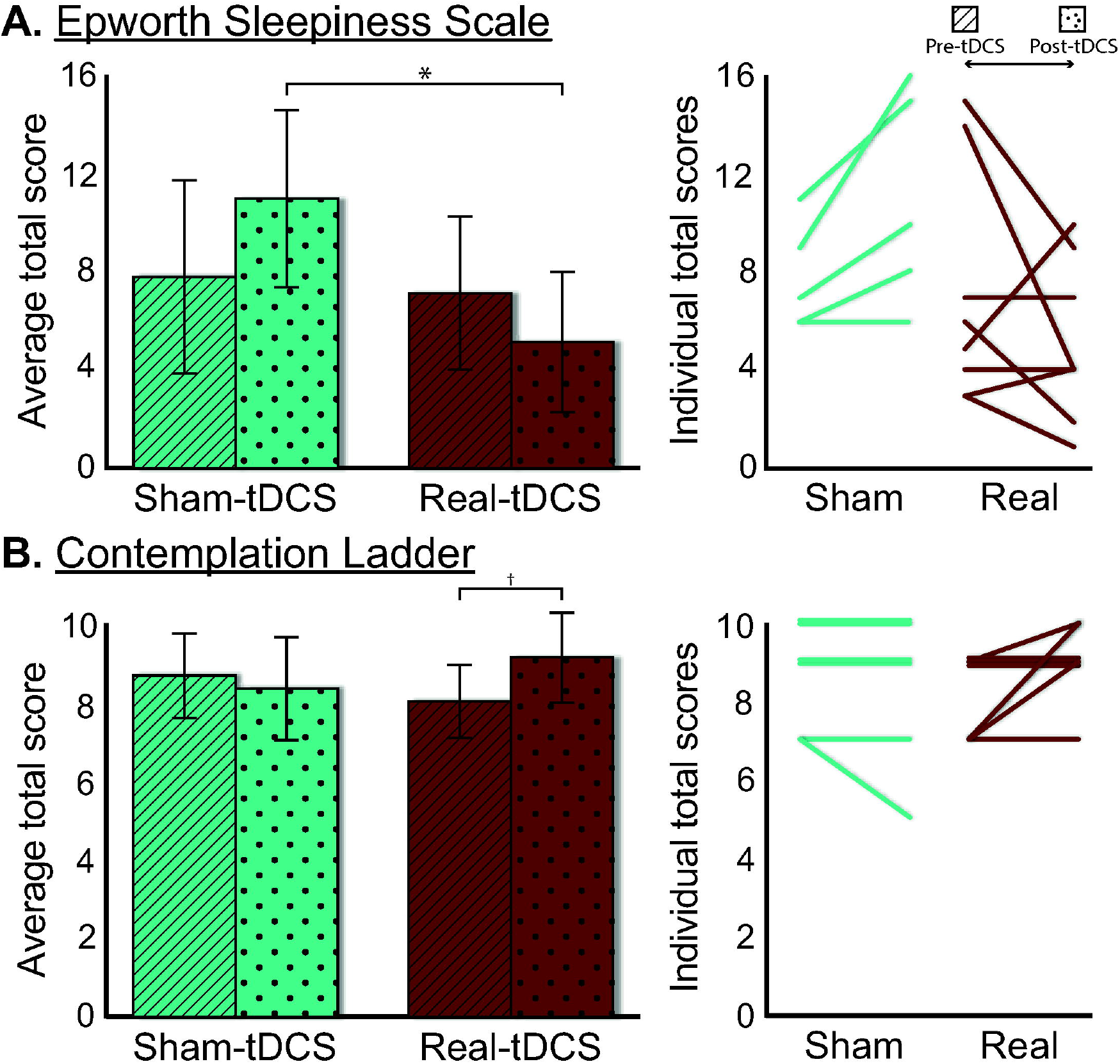
Significant Group × Time interactions for sleepiness and motivation to change. A) Left panel: group-averaged total score on the Epworth Sleepiness Scale at both study timepoints (pre- and post-tDCS). The significant Group × Time interaction was driven by a group difference post-tDCS (* p < 0.05). Right panel: Individual scores showing change from pre- to post-tDCS in each group. B) Left panel: group-averaged total score on the Contemplation Ladder at both study timepoints (pre- and post-tDCS). The significant Group × Time interaction was driven by a trend for an increase in readiness to change in the real-tDCS group only (^**†**^ p = 0.05). Right panel: Individual scores showing change from pre- to post-tDCS in each group.

**Figure 4:**
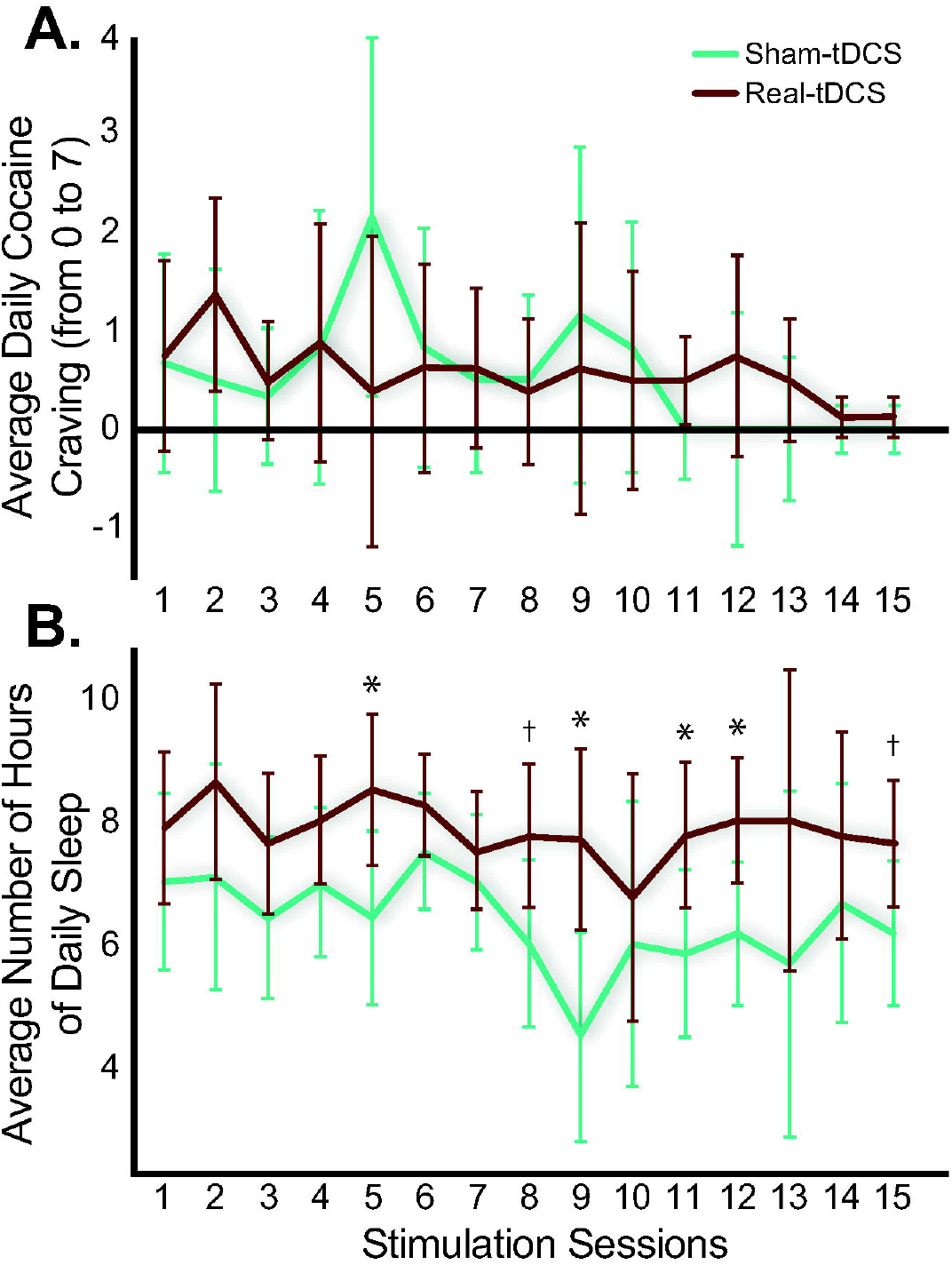
Group-averaged number of hours of daily sleep and daily self-reported cocaine craving for all stimulation sessions. A) A trend for a Time main effect (p = 0.075) was found for the daily self-reported cocaine craving on a scale of 0 to 7. A linear contrast reached significance (p < 0.05) suggesting a linear decrease in craving over the 15 sessions across all subjects. B). A Group main effect (Real- > Sham-tDCS; p < 0.05) was driven by significantly more sleeping hours after the 5^th^, 9^th^, 11^th^, and 12^th^ sessions with similar trends for the 8^th^ and 15^th^ sessions. ^**†**^ p = 0.05; * p < 0.05.

### Secondary Outcome Measures

The 2 × 2 ANOVAs with repeated measures showed a significant Group × Time interaction for the Epworth Sleepiness Scale [F(1,11) = 5.18, p = 0.044, *d =* 1.4, Figure 3A]; the main effects of Group and Time were not significant [F(1,11) < 3.05, p > 0.109, d < 1.1]. Follow-up analyses showed that this interaction was explained by a group difference (real-tDCS < sham-tDCS) post [t(12) = 2.48, p = 0.029] but not pre-tDCS [t(11) = 0.30, p = 0.774)] suggesting that individuals with CUD who received real-tDCS reported less sleepiness than those in the sham-tDCS group after all 15 tDCS sessions. The 2 × 15 ANOVA of the daily reports of the amount of sleep yielded corroborating results (Figure 4B). Here, the Group by Time interaction did not reach significance [F(14,168) = 0.74, p = 0.729, d = .13], instead revealing a Group main effect (real-tDCS > sham-tDCS) [F(1,12) = 7.15, p = 0.020, d = 1.5]. Subsequent exploratory t-tests however showed real- as compared to the sham-tDCS participants to report significantly more hours of sleep after the 5^th^ [t(12) = −2.43, p = 0.032], 9^th^ [t(12) = −3.08, p = 0.009], 11^th^ [t(12) = −2.31, p = 0.040], and 12^th^ [t(12) = −2.58, p = 0.024] sessions with similar trends for the 8^th^ [t(12) = −2.41, p = 0.054] and 15^th^ [t(12) = −2.03, p = 0.065] sessions but not for the first 4 sessions [t(12) < −1.52, p > 0.155], indicating that the groups started differentiating later during treatment (See Figure 4B). In contrast, sleep quality, as measured by the Pittsburgh Sleep Quality Index, showed a Time main effect, indicative of a decrease in the overall sleep quality during inpatient treatment in both groups [F(1,11) = 5.02, p = 0.047, *d =* 1.4]. A Group × Time interaction [F(1,12) = 5.40, p = 0.039, *d =* 1.3, Figure 3B] was also significant for the Contemplation Ladder where simple effects showed a trend towards increase in readiness to change in the real-tDCS [t(7) = −2.35, p = 0.051] but not sham-tDCS [t(5) = 1.0, p = 0.363] group (Figure 3B), suggesting higher readiness to change after the stimulation sessions in the real-tDCS group.

### One-Month Follow-Up

Using 2 (Group) × 3 (Time) ANOVAs with repeated measures, analyses were carried out for the three outcome variables that showed significant Group × Time interactions (or a trend) and for all outcome variables (in addition to the craving intensity item on the Cocaine Selective Severity Assessment) that showed a significant Time effect (Table 1, Figure 5). While correcting for baseline value, no Group × Time interaction was found for the 5-item OCCS craving subscale [F(2,18) = 2.32, p = 0.127, Figure 5A] but a significant Group effect is otherwise found where the real-tDCS group shows lower craving scores than the sham-tDCS group [F(1,9) = 6.31, p < 0.033]. Although no significant effect was found for the 5-item Cocaine Craving Questionnaire [F(2,20) < 2.11, p > 0.147, Figure 5B] either, the craving question of the Cocaine Selective Severity Assessment showed a significant Time effect, driven by decreases in cocaine cravings during treatment and their increase back to baseline one month following the protocol [F(2,20) = 3.62, p = 0.045]. A significant interaction effect for the Epworth Sleepiness Scale [F(2,9) = 6.07, p = 0.011, Table 2 and Figure 5C] was driven by a significant time effect only in the real-tDCS group [F(1,6) = 5.71, p = 0.021] such that participants reported more sleepiness at follow-up when compared to the immediate post-tDCS assessment (p = 0.009) suggesting that values returned to baseline after one month follow-up in this group. There were no significant effects for the Contemplation Ladder [F(2,20) < 1.75, p > 0.199, Figure 5D] nor for the Pittsburgh Sleep Quality Index [F(2,18) < 2.20, p > 0.172, Figure 5F]. The environment domain of the Quality of Life Questionnaire showed a significant Time effect [F(2,20) = 5.70, p = 0.011, Figure 5E] where values were higher in both groups after the 15 tDCS sessions, remaining elevated after a one-month follow-up (p < 0.05).

**Table 2.**
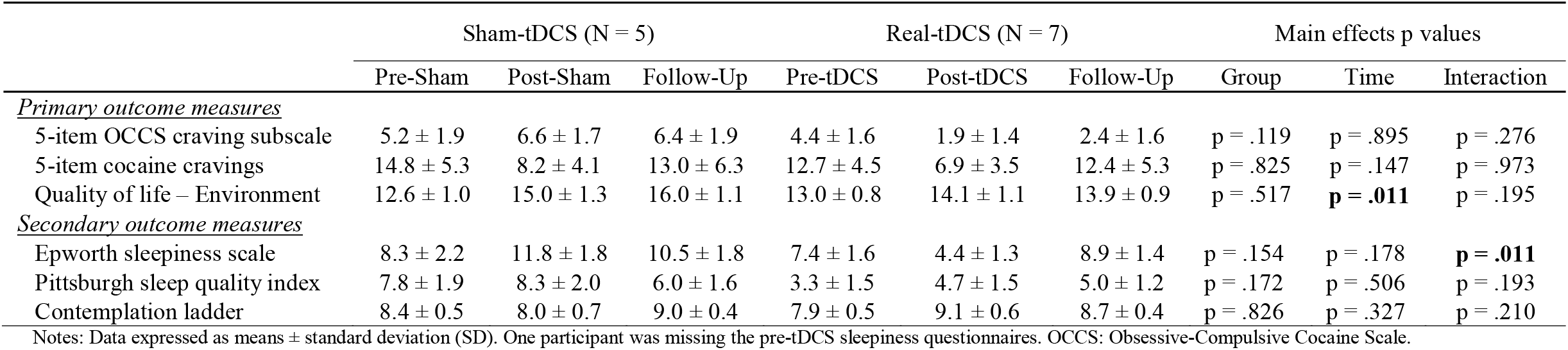
Follow-up analyses of outcome measures showing a significant effect of tDCS treatment (Time or Time by Group) in sham- and real-tDCS groups

|  | Sham-tDCS (N = 5) |  |  | Real-tDCS (N = 7) |  |  | Main effects p values |  |  |
| --- | --- | --- | --- | --- | --- | --- | --- | --- | --- |
|  | Pre-Sham | Post-Sham | Follow-Up | Pre-tDCS | Post-tDCS | Follow-Up | Group | Time | Interaction |
| *<u>Primary outcome measures</u>* |  |  |  |  |  |  |  |  |  |
| 5-item OCCS craving subscale | 5.2 ± 1.9 | 6.6 ± 1.7 | 6.4 ± 1.9 | 4.4 ± 1.6 | 1.9 ± 1.4 | 2.4 ± 1.6 | p = .119 | p = .895 | p = .276 |
| 5-item cocaine cravings | 14.8 ± 5.3 | 8.2 ± 4.1 | 13.0 ± 6.3 | 12.7 ± 4.5 | 6.9 ± 3.5 | 12.4 ± 5.3 | p = .825 | p = .147 | p = .973 |
| Quality of life – Environment | 12.6 ± 1.0 | 15.0 ± 1.3 | 16.0 ± 1.1 | 13.0 ± 0.8 | 14.1 ± 1.1 | 13.9 ± 0.9 | p = .517 | **p = .011** | p = .195 |
| *<u>Secondary outcome measures</u>* |  |  |  |  |  |  |  |  |  |
| Epworth sleepiness scale | 8.3 ± 2.2 | 11.8 ± 1.8 | 10.5 ± 1.8 | 7.4 ± 1.6 | 4.4 ± 1.3 | 8.9 ± 1.4 | p = .154 | p = .178 | **p = .011** |
| Pittsburgh sleep quality index | 7.8 ± 1.9 | 8.3 ± 2.0 | 6.0 ± 1.6 | 3.3 ± 1.5 | 4.7 ± 1.5 | 5.0 ± 1.2 | p = .172 | p = .506 | p = .193 |
| Contemplation ladder | 8.4 ± 0.5 | 8.0 ± 0.7 | 9.0 ± 0.4 | 7.9 ± 0.5 | 9.1 ± 0.6 | 8.7 ± 0.4 | p = .826 | p = .327 | p = .210 |

Notes: Data expressed as means ± standard deviation (SD). One participant was missing the pre-tDCS sleepiness questionnaires. OCCS: Obsessive-Compulsive Cocaine Scale.

**Figure 5:**
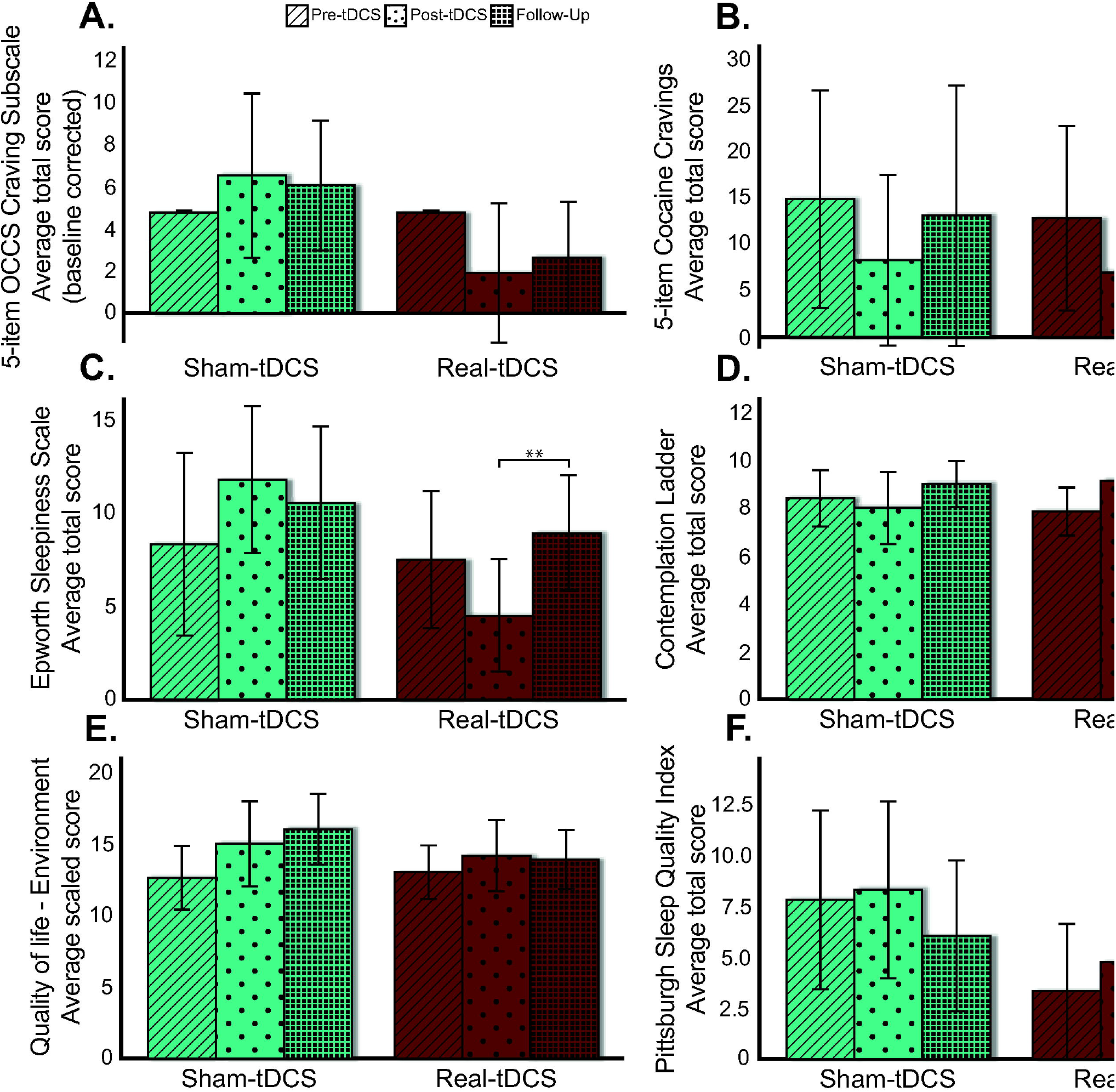
One-month follow-up of craving, daytime sleepiness, readiness to change, sleep quality, and quality of life. Group-averaged total scores on all measures that showed significant Group × Time interactions and for all outcome variables that showed a significant Time effect (or a trend) for all three study timepoints (pre-tDCS, post-tDCS, and follow-up). Those measures were the 5-item OCCS craving subscale (A), 5-item Cocaine Cravings Questionnaire (B), Epworth Sleepiness Scale (C), Contemplation Ladder (D), the Environment domain of the abbreviated version of the World Health Organization Quality of Life questionnaire (E), and the Pittsburgh Sleep Quality Index (F). A Group × Time interaction on the Epworth Sleepiness Scale was driven by a time effect only in the real-tDCS group such that participants reported more sleepiness at follow-up when compared to the immediate post-tDCS assessment (** p < 0.01). A significant Group effect was found for the 5-item OCCS craving subscale when correcting for baseline values where the real-tDCS group showed lower cravings than the sham-tDCS group (p < 0.05). A significant Time effect was also found for the Environment domain of the Quality of Life questionnaire (p < 0.05).

### STRAP-R

Our 5-way mixed ANOVA showed main effects of Group [sham-tDCS > real-tDCS, F(1,12) = 9.48, p = 0.010], Reward [Food = Sex > Drug, F(2,24) = 16.35, p < 0.0001], and Question [“liking” > “wanting”, F(1,12) = 9.24, p = 0.010), and Situation × Question [F(2,24) = 10.22, p = 0.001] and Reward × Situation [F(4,48) = 16.57, p < 0.0001] interactions quantified by a significant 3-way Reward × Situation × Question interaction [F(4,48) = 4.04, p = 0.007] (See Table 3). To follow up on the significant interactions, 3 (Situation) × 2 (Question) ANOVAs with repeated measures were carried out independently for each Reward type. For the Food reward, there were significant Situation [“now” = “in general” > “under the influence”, F(1.27,16.45) = 6.54, p < 0.016] and Question effects [“liking” > “wanting”, F(1,13) = 5.17, p = 0.041]. The Sex reward showed the same Question effect [“liking” > “wanting”, F(1,13) = 5.56, p = 0.035]; there were no other significant effects [F < 1.40, p > 0.269]. Consistent with our prior reports (Goldstein *et al.*, 2010), for the Drug reward, there was a significant Situation main effect [“now” < “in general” < “under the influence”, F(2,26) = 33.56, p < 0.0001]. A significant Situation × Question interaction effect was also observed [F(1.39, 18.03) = 13.02, p = 0.001] and showed that “liking” was higher than “wanting” for both the “now” (t(13) = 3.37, p = 0.005) and “in general” (t(13) = 2.68, p = 0.019) situations whereas “wanting” was higher than “liking” for the “under the influence” (t(13) = −3.30, p = 0.006) situation. No significant effects for Time [F(1,12) = 0.44, p = 0.522] or interactions with Time or Group [F < 2.18, p > 0.135] were however found for the STRAP-R questionnaire.

**Table 3.** Sensitivity to Reinforcement of Addictive and other Primary Rewards (STRAP-R) measures in sham- and real-tDCS groups.

| Reward | Situation | Question | Sham-tDCS (N = 6) |  | Real-tDCS (N = 8) |  |
| --- | --- | --- | --- | --- | --- | --- |
|  |  |  | Pre-sham | Post-sham | Pre-tDCS | Post-tDCS |
| <u>Food</u> | Currently | Liking | 4.5 ± 0.5 | 4.7 ± 0.8 | 3.9 ± 1.6 | 4.4 ± 0.9 |
|  |  | Wanting | 3.8 ± 1.5 | 4.3 ± 1.2 | 3.8 ± 1.6 | 3.4 ± 1.4 |
|  | In General | Liking | 4.3 ± 0.8 | 5.0 ± 0.0 | 4.0 ± 0.9 | 4.3 ± 0.9 |
|  |  | Wanting | 3.2 ± 0.4 | 4.7 ± 0.5 | 4.0 ± 0.9 | 4.3 ± 1.2 |
|  | Under the influence/high | Liking | 2.7 ± 1.9 | 4.0 ± 1.7 | 2.9 ± 1.9 | 2.9 ± 1.8 |
|  |  | Wanting | 2.7 ± 1.9 | 3.2 ± 1.6 | 3.1 ± 2.0 | 2.9 ± 2.0 |
| <u>Sexual Activity</u> | Currently | Liking | 4.8 ± 0.4 | 5.0 ± 0.0 | 3.8 ± 1.2 | 3.8 ± 1.8 |
|  |  | Wanting | 4.5 ± 1.2 | 4.8 ± 0.4 | 3.3 ± 1.6 | 3.5 ± 1.9 |
|  | In General | Liking | 4.8 ± 0.4 | 4.8 ± 0.4 | 4.0 ± 1.1 | 4.1 ± 1.5 |
|  |  | Wanting | 4.7 ± 0.8 | 4.7 ± 0.8 | 3.3 ± 1.4 | 3.4 ± 1.5 |
|  | Under the influence/high | Liking | 4.7 ± 0.8 | 3.8 ± 1.6 | 2.9 ± 1.6 | 3.6 ± 1.8 |
|  |  | Wanting | 4.7 ± 0.8 | 4.0 ± 1.5 | 2.5 ± 1.3 | 3.6 ± 1.7 |
| <u>Drug</u> | Currently | Liking | 2.0 ± 1.7 | 2.7 ± 1.6 | 2.1 ± 1.4 | 2.0 ± 1.6 |
|  |  | Wanting | 1.5 ± 0.8 | 1.2 ± 0.4 | 1.4 ± 0.5 | 1.4 ± 0.7 |
|  | In General | Liking | 3.0 ± 1.3 | 2.2 ± 1.6 | 3.1 ± 1.6 | 2.6 ± 1.8 |
|  |  | Wanting | 2.5 ± 1.8 | 1.5 ± 0.8 | 1.5 ± 0.5 | 1.9 ± 1.5 |
|  | Under the influence/high | Liking | 3.8 ± 1.0 | 2.7 ± 1.2 | 2.8 ± 1.6 | 3.0 ± 1.5 |
|  |  | Wanting | 4.3 ± 0.8 | 4.0 ± 1.3 | 3.6 ± 1.8 | 4.0 ± 1.4 |
| **Significant Statistical Effects** |  |  |  |  |  |  |
| <u>Reward Effect</u><br>(p < 0.0001) |  | <u>Question Effect</u><br>(p < 0.01) | ----- <u>Group Effect</u> -----<br>(p < 0.01) |  |  |  |
| <u>Reward × Situation Interaction</u><br>(p < 0.0001) |  |  |  |  |  |  |
| <u>Situation × Question Interaction</u><br>(p < 0.001) |  |  |  |  |  |  |
| <u>Reward × Situation × Question Interaction</u><br>(p < 0.01) |  |  |  |  |  |  |

Notes: Data expressed as means ± standard deviation (SD). STRAP-R values represent the rating on a Likert scale ranging from “Somewhat” (1) to “Extremely (5).”

## Discussion

The main goal of this Phase 1 clinical trial was to replicate previous findings showing a reduction in subjective cocaine cravings associated with repeated tDCS over the dlPFC in individuals with CUD living in an inpatient drug-rehabilitation facility (Batista *et al.*, 2015). Using a portable device in such a non-medical setting, our study also aimed at assessing the feasibility of an extended protocol of 15 repeated sessions of tDCS. Feasibility was clearly evident in the completion of all 15 sessions in 14 of the 17 subjects who started this study (82% retention rate); future studies need to take into account drop-outs driven by subjects’ transfer to different treatment facilities that may not allow participation in research-related activities. Beyond treatment-as-usual effects evident in longer abstinence duration, enhanced perceived quality of the environment and sleep measures, and in reduced impulsivity, our main finding provided partial support for the impact of real-tDCS on reducing craving. Importantly, we observed decreased self-reported daytime sleepiness and increased readiness to change in the real-tDCS group only. The real-tDCS effect on sleepiness was not maintained after a one-month follow-up, a result that needs to be tested with larger samples.

Previous findings showed a significant decrease in subjective cocaine craving specifically in the real-tDCs group (Batista *et al.*, 2015), bolstered by similar results in individuals using other psychostimulants such as methamphetamine and nicotine (Fecteau *et al.*, 2014; Shahbabaie *et al.*, 2014, 2018; Hajloo *et al.*, 2019; Martinotti *et al.*, 2019; Alizadehgoradel *et al.*, 2020). Our results provide partial support for our main hypothesis for a similar unique effect of real-tDCS on craving in our subjects. Using the 5-item Obsessive-Compulsive Cocaine Scale craving subscale (De Wildt *et al.*, 2005), the tool used by the previous study, a Group by Time interaction showed a trend toward significance when correcting for baseline values and using a one-tailed threshold. For this effect to reach significance, 30 individuals with CUD (15 in each group) would need to be enrolled in a Phase 2 study. Our other craving measures mainly showed a Time main effect such that participants in both groups reported less cravings after vs. before the experiment. This Time effect was observed for the self-reported craving intensity item on the Cocaine Selective Severity Assessment (Kampman *et al.*, 1998) supported by a significant linear decrease over time in the 15 daily self-reports of cocaine craving with a similar trend also for the 5-item Cocaine Craving Questionnaire (Tiffany *et al.*, 1993). Overall, this decrease in craving for both the study groups can be attributed to drawing our sample from treatment-seeking individuals living in an inpatient facility, a controlled and structured therapeutic environment. Batista and colleagues (2015) also reported that only the real-tDCS (and not sham) treatment was associated with a decrease in anxiety and depressive symptoms and higher levels in self-reported quality of life (in the psychological and environment domains). Consistent with the treatment environment, we observed a significant increase in the perception of the quality of the environment in both the real- and sham-tDCS groups, while effects for anxiety and depression were not significant, potentially because of insufficient statistical power and the need for larger sample sizes. Our results also showed a significant decrease in impulsivity with time in inpatient treatment, with both groups better able to value monetary reinforcement even if delayed in time (Odum, 2011; Mejía-Cruz *et al.*, 2016). The association between impulsivity and craving deserves further exploration especially in light of a recent study in patients with alcohol use disorder showing that increased impulsivity was associated with higher craving, which, in turn, increased the likelihood of alcohol use in treatment, suggesting that craving mediates the relationship between impulsivity and the likelihood to relapse (Coates *et al.*, 2020).

Our exploratory results showed a specific decrease in self-reported daytime sleepiness, suggesting an increase in alertness that is characteristic of initial abstinence (Conroy & Arnedt, 2014), only in the real-tDCS (and not sham) group. Although the Group by Time interaction did not reach significance in this small sample, the daily measures of sleep quantity showed similar effects such that the real-tDCS group reported sleeping on average more hours than the sham-tDCS group, an effect reaching significance in later sessions only (starting at the fifth session), suggesting a cumulative effect of real-tDCS on sleep quantity. The seemingly contradictory significant Time effect on the Pittsburgh Sleep Quality Index, where both groups reported a decrease in sleep quality after all tDCS sessions, could reflect the inpatient conditions where subjects share bedrooms similar to dormitories with no possibility for individual adjustments (e.g., in sleep/wake cycle). Overall, our results are consistent with other tDCS studies targeting the dlPFC with different electrode setups (left dlPFC anodal electrode and single or multiple cathodal electrodes throughout the scalp) where real-tDCS increased the total sleep duration in normal adults (Frase *et al.*, 2016) and older healthy adults (Sheng *et al.*, 2018); our results are not consistent with studies reporting increases in sleep-quality, e.g., in euthymic bipolar patients (Minichino *et al.*, 2014). The exact mechanisms underlying the putative effect of tDCS on sleep are yet to be fully investigated but of note is an alteration of resting-state functional MRI connectivity between the default mode network and subcortical regions, including the amygdala, hippocampus, striatum, globus pallidus, and thalamus, after high-definition tDCS in older adults (Sheng *et al.*, 2018), known for their crucial role in sleep-wake functions (Pace-Schott & Hobson, 2002; Sämann *et al.*, 2011; De Havas *et al.*, 2012).

In addition, participants in the real-tDCS but not sham-tDCS group reported a higher motivation to change as measured by the Contemplation Ladder after (vs. before) all 15 sessions. Motivation to change is an inherent part of drug rehabilitation, associated with engagement in recovery-oriented behaviors (Gregoire & Burke, 2004) and predictive of changes in drug consumption (Heather *et al.*, 1993). During a recent functional MRI reappraisal task performed by cocaine addicted individual, readiness to change was positively correlated with the right dlPFC response to emotional regulation and negatively with its response to emotional processing, prompting the authors to propose that a greater top-down control of negative emotionality and reactivity is associated with maintaining goal-oriented behaviors (Contreras-Rodríguez *et al.*, 2020). It is therefore possible that our tDCS protocol that targeted the right dlPFC enhanced subjects’ motivation to change through a top-down executive control mechanism, potentially contributing to both the persistence towards reducing drug use and the ability to flexibly learn alternatives to current strategies for maintaining abstinence, consistent with the iRISA model (Volkow *et al.*, 2010; Goldstein & Volkow, 2011; Hobkirk *et al.*, 2019) as remains to be studied with objective measures.

This iRISA syndrome is associated with enhanced attribution of salience to drug-related cues and with decreased sensitivity to non-drug reinforcers; we therefore evaluated the effects of tDCS on self-reported sensitivity to reinforcement of addictive and other primary rewards using the STRAP-R questionnaire. Although we did not observe significant Time or Group interactions, suggesting this sensitivity may mostly reflect trait measures not affected by tDCS, we corroborated previous results where the subjective value of drug and non-drug reinforcers depended on the situation (Goldstein *et al.*, 2010). More precisely, individuals with CUD gave a higher “liking” than “wanting” ratings in all situations (“current”, “in general” and “under the influence”) for non-drug reinforcers (food and sex) whereas they exhibited more drug “wanting” than “liking” when reporting about being “under the influence”. These results support the idea that drug stimuli are wanted (potentially underlying habit and compulsion) more than they are liked (hedonic properties) especially during a drug-cue context in addiction (Goldstein & Volkow, 2002, 2011; Goldstein *et al.*, 2010).

A major limitation of this study is its small sample size (especially during the follow-up assessment), potentially reducing statistical power by increased interindividual variability characterizing the effects of non-invasive brain stimulation on cortical plasticity (Ridding & Ziemann, 2010; López-Alonso *et al.*, 2014). Another limitation is the use of self-report to assess craving, which could reflect habituation and/or floor or other effects (e.g., compromised self-awareness). A Phase 2 clinical trial should include more participants in each group as well as objective measures of cocaine craving such as drug-related cue-reactivity (e.g., assessed by electroencephalography and event-related potentials), which may be more sensitive to our effects of interest. Indeed, these objective measures tracked cue-induced incubation of craving, showing a discrepancy with subjective measures of cocaine craving during abstinence in individuals with CUD (Parvaz *et al.*, 2016). Our study suggests only a transient effect of tDCS on sleepiness and craving when tested about one month after the last tDCS session. Further studies are needed to establish the tDCS dose and duration needed for optimally maintaining any beneficial impact in inpatients with CUD. Finally, the impact of medications (taken by all our participants) on results remains to be systematically studied. Some studies suggest that pharmacotherapy in combination with electrical brain stimulation yield greater effects (than brain stimulation or pharmacotherapy alone) in depression (Brunoni *et al.*, 2013, 2014), although other studies suggest no significant effects of pharmacotherapy on decreasing craving, observed in both the experimental and control groups after 10 sessions of repeated tDCS in inpatients with CUD (Klauss *et al.*, 2018).

To summarize, our study in individuals with CUD showed a trend towards a unique effect of real-tDCS on craving, an effect that could reach significance with 15 subjects per group. In this small sample, inpatient treatment itself, and the ensuing longer abstinence, was associated with a reduction in self-reported craving and improvements in the perceived quality of the environment as well as with reduced impulsivity/improvements in valuation of delayed rewards (but not with changes in the value attributed to drug-related rewards especially when thinking about a drug-relevant context). Interestingly, real-tDCS (but not sham-tDCS) was associated with reductions in daytime sleepiness and increases in readiness to change potentially suggesting the fortification of executive cognitive functions supported by the dlPFC. A main result from this Phase 1 project was also the demonstration of the feasibility of using a portable tDCS device 15 times over 5 weeks in inpatients with CUD. Overall, our results suggest the need for extending this proof-of-concept study to establish the use of tDCS as an add-on therapy in drug addiction. Specifically, these results call for a larger sample further incorporating objective (e.g., psychophysiological) outcome measures and ultimately assessing the remote and self-administration of tDCS (e.g., in other in- and out-patient drug rehabilitation facilities, in combination with other cognitive and pharmacological therapies, and ultimately at home) for the purpose of managing drug addiction symptomatology.

## Data Availability

Data acquired for this project are available upon request to Dr. Rita Z. Goldstein.

## Acknowledgements

This study was supported by NIDA 271201800035C-0-0-1 “tDCS to reduce craving in cocaine addiction” (SBIR Phase I), by R01DA041528 to Dr. Goldstein, and by the Canadian Institutes of Health Research (Postdoctoral research award to Dr. Gaudreault).

## Author Contributions

<u>Study design</u>: Datta, Parvaz, Parra, Alia-Klein, Goldstein and Nakamura-Palacios (consultant). <u>Intervention blinding</u>: Parvaz and Goldstein. <u>Study implementation</u>: Gaudreault, Datta, Malaker, and Wagner. <u>Data acquisition</u>: Gaudreault, Sharma, King, Malaker, Wagner and Vasa. <u>Data analysis</u>: Gaudreault and Sharma. <u>Manuscript writing</u>: Gaudreault, Sharma and Goldstein. <u>Manuscript review</u>: Datta, Nakamura-Palacios, Parvaz, Parra, Alia-Klein and Goldstein.

## Data Accessibility

Data acquired for this project are available upon request to Dr. Rita Z. Goldstein.

